# Seroprevalence and Risk Factors of SARS CoV-2 in Health Care Workers of Tertiary-Care Hospitals in the Province of Khyber Pakhtunkhwa, Pakistan

**DOI:** 10.1101/2020.09.29.20203125

**Authors:** Mohsina Haq, Asif Rehman, Muhammad Noor, Jawad Ahmad, Junaid Ahmad, Muhammad Irfan, Saeed Anwar, Sajjad Ahmad, Said Amin, Fawad Rahim, Najib Ul Haq

## Abstract

**Background:** High number of SARS-CoV-2 infected patients has overburdened healthcare delivery system, particularly in low-income countries. In the recent past many studies from the developed countries have been published on the prevalence of SARS CoV-2 antibodies and the risk factors of COVID-19 in healthcare-workers but little is known from developing countries.

**Methods:** This cross-sectional study was conducted on prevalence of SARS-CoV-2 antibody and risk factors for seropositivity in HCWs in tertiary-care hospitals of Peshawar city, Khyber Pakhtunkhwa province Pakistan.

**Results:** The overall seroprevalence of SARS CoV-2 antibodies was 30·7% (CI, 27·8–33·6) in 1011 HCWs. Laboratory technicians had the highest seropositivity (50·0%, CI, 31·8–68·1).

Risk analysis revealed that wearing face-mask and observing social-distancing within a family could reduce the risk (OR:0·67. p<0·05) and (OR:0·73. p<0·05) while the odds of seropositivity were higher among those attending funeral and visiting local-markets (OR:1·83. p<0·05) and (OR:1·66. p<0·01). In Univariable analysis, being a nursing staff and a paramedical staff led to higher risk of seropositivity (OR:1.58. p< 0·05), (OR:1·79. p< 0·05). Fever (OR:2·36, CI, 1·52– 3·68) and loss of smell (OR:2·95, CI: 1·46–5·98) were significantly associated with increased risk of seropositivity (p<0.01). Among the seropositive HCWs, 165 (53·2%) had no symptoms at all while 145 (46·8%) had one or more symptoms.

**Conclusion:** The high prevalence of SARS-CoV-2 antibodies in HCWs warrants for better training and use of protective measure to reduce their risk. Early detection of asymptomatic HCWs may be of special importance because they are likely to be potential threat to others during the active phase of viremia.

## Introduction

The first case of Corona Virus was reported in BMJ in 1965^1^ Many corona viruses have been recovered from animals or humans, however, only two of them have gained attention in the past two decades.^2,3,4^ The transmission of virus to others is typically like that of the “common cold”. Healthcare workers are exposed to and at higher risk of acquiring infection while dealing with patients suffering from highly infectious diseases like COVID-19. PCR may be negative even in acute phase in certain cases.^5^ Antibodies tests (anti SARS-CoV-2 antibodies) may be useful in diagnosing PCR negative cases and also provide information about past infection.^5,6^ A Cochrane review of 54 studies on antibody testing reported that 94% patients may be positive after the third week of onset of symptoms^7^ and hence may be a better index of past exposure to SARS CoV-2. The role of antibodies in preventing further infection from COVIC-19 is still not clear^8^ however it is assumed that antibodies may provide some protection.^9^

Many studies have been published on the prevalence of SARS CoV-2 antibodies in healthcare workers from developed countries in recent past, however little is known from developing countries. To our knowledge this is the first study of assessing SARS-CoV-2 antibodies of HCWs form both public and private tertiary care hospitals in Peshawar, Pakistan. The present study aims to estimate the seroprevalence of SARS-CoV-2 in HCWEs and explores the possible risk factors of exposure to SARS-CoV-2.

## Methods

### Study design and participants

This is a cross-sectional study, following the STROBE (Strengthening the Reporting of Observational Studies in Epidemiology) reporting guidelines, conducted from June 15 to 29, 2020 using purposive sampling technique. The number of HCWs included in the study was 1011. to participate, were included in the study. The HCWS included doctors, paramedics, nurses, medical technicians, laboratory and other staff of the hospitals.

### Data Collection procedure

The study was approved by Institutional Review Board of Prime Foundation Pakistan. Data about detailed history of risk factors, co-morbid factors, demographic information and symptoms was collected on a semi-structured proforma. Five ml peripheral venous blood was collected in Li Heparinised tube, after informed, serum separated using 2500 rpm centrifuge and stored in labelled serum cup for analysis using 20 micro litre serum volume while remaining serum was stored at – 80 C^0^ temperature. COBAS e411 system was used for Immunoassay.

### Detection of SARS CoV-2 antibodies

The FDA approved kit was used for detection of Anti-SARS-CoV-2 antibodies which has high specificity (100% and sensitivity (more than 98·8%) according to the manufacturers.^10^, however Public Health England estimated its specificity to be 100% but a sensitivity of 87%.^11^ Results were interpreted against a cut off value of 1 AU/ml and less than 1 AU/ml was considered Negative and more than or equal to 1AU/ml as positive.

### Statistical Analysis

**S**tatistical analyses were performed using SPSS v.24·0. The means and standard deviations were used to present the continuous variables and the categorical variables were described as the counts and the percentages. Variables with *p* values < 0·01 in the univariate analysis were further used for a multivariate logistic regression analysis and *p* value ≤0·05 was considered significant.

## Results

### Socio-demographic characteristics

The demographic characteristics of healthcare workers are summarized in table 1 below. The FCWS included 688 (68·1%) males and 323 (31·9%) female. The mean age was 33.6 years (SD ±10·5) while 454 (45·0%) were in the age group 20–29 years and 312 (31.0%) 30–39 years. and only 34 (3·40%) in age group 60 years and above.

**Table 1:** Sociodemographic Characteristics

|  | Number of<br>participants<br>*N* | Seropositive<br>*N (%)* | Seronegative<br>*N (%)* | Seroprevalence (95% CI)<br>Binomial Exact |
| --- | --- | --- | --- | --- |
| **Overall** | 1011 | 310 (30.7%) | 701 (69.3%) | 30.7% (27.8-33.6) |
| **Gender** |  |  |  |  |
| Male | 688 | 219 (31.8%) | 469 (68.2%) | 31.8% (28.3 – 35.4) |
| Female | 323 | 91 (28.2%) | 232 (71.8%) | 28.2% (23.3 – 33.4) |
| **Age groups** |  |  |  |  |
| 20-29 | 532 | 157 (29.5%) | 375 (70.5%) | 29.5% (25.7 – 33.5) |
| 30-39 | 266 | 90 (33.8%) | 176 (66.2%) | 33.3% (28.1 – 39.8) |
| 40-59 | 127 | 40 (31.5%) | 87 (68.5%) | 31.5% (23.5 – 40.3) |
| 50-59 | 62 | 22 (35.5%) | 40 (64.5%) | 35.5% (23.7 – 48.6) |
| 60 and above | 24 | 1 (4.2%) | 23 (95.8%) | 4.2% (0.1 – 2.11) |
| **Professional**<br>**category** |  |  |  |  |
| Consultant | 154 | 28 (18.2%) | 126 (81.8%) | 18.2% (12.4 – 25.1) |
| MO/TMO | 181 | 36 (19.9%) | 145 (80.1%) | 19.9% (14.3 – 26.4) |
| HO | 114 | 21 (18.4%) | 93 (81.6%) | 18.4% (11.7 – 26.7) |
| Nursing Staff | 209 | 81 (38.8%) | 128 (61.2%) | 38.8% (32.1 – 45.7) |
| Paramedical<br>Staff | 157 | 66 (42.0%) | 91 (58.0%) | 42.0% (34.2 – 50.1) |
| Ward Staff | 88 | 35 (39.8%) | 53 (60.2%) | 39.8% (29.4 – 50.7) |
| Lab Tec | 32 | 16 (50.0%) | 16 (50.0%) | 50.0% (31.8 – 68.1) |
| Other | 76 | 27 (35.5%) | 49 (64.5%) | 35.5% (24.8 – 47.3) |
| **Symptom**<br>**Status** |  |  |  |  |
| Symptomatic | 457 | 145 (31.7%) | 312 (68.3%) | 31.7% (27.4 – 36.2) |
| Asymptomatic | 554 | 165 (29.8%) | 389 (70.2%) | 29.8% (26.0 – 33.7) |

The professional categories of HCWs were, nursing staff (26·1%), paramedical staff (21·3%), trainee doctors / medical officers (11·6%), ward staffs (11·3%), consultants (9%), house officers (6.8%), Lab Technicians 5·2% and 8·7% were ward support staff members.

### Seroprevalence of antibodies against SARS CoV-2

The overall seroprevalence of SARS-CoV-2 antibodies was 30·7% (CI 95%: 27·8 – 33·6). The seroprevalence was not significantly different (P>0·02) in males 31·8% (CI 95%: 28·3 – 35·4) than female 28·2% (CI 95%: 23·3 – 33·4) female subjects [Table: 1].

The age wise seroprevalence of SARS-CoV-2 antibodies was 29·5% (95% CI 25·7–33·5) in age group 20-29 years, 33·3%(95% CI, 28·1 – 39·8) and it increasing with older age until plateauing at age group 50-59 35·5% (95% CI, 23·7 – 48·6) while it declined in group 60 years and above (4·2%, 95% CI, 0.1 – 2·1) [Table 1 & Graph 1].

**Graph 1:**
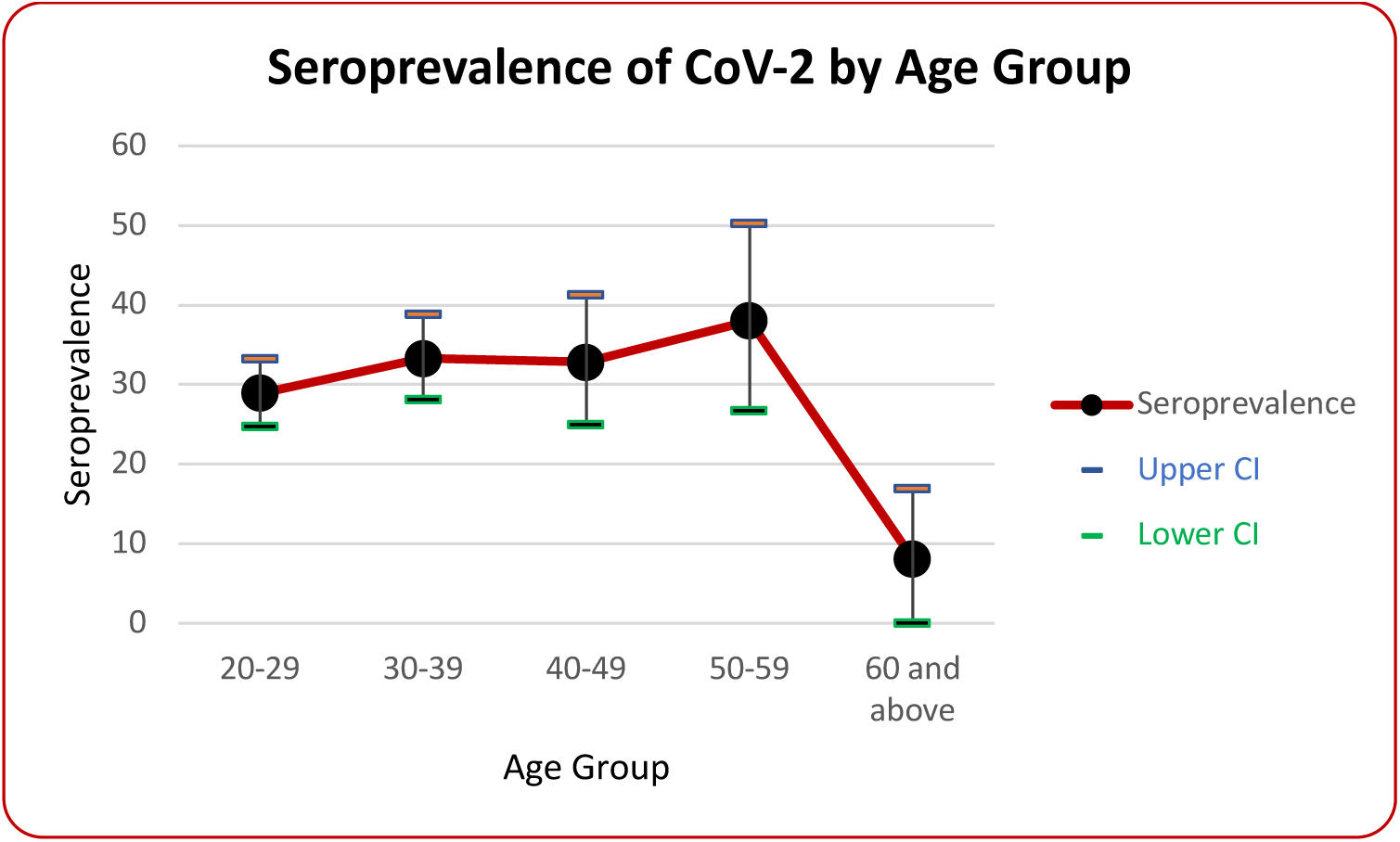

In different professional category, the highest seroprevalence were identified in Lab technicians (50·0%, 95% CI 31·8–68·1) followed by paramedical staff (42.0%, 95% CI 34.2 – 50.1), ward staff (39·8%, 95% CI 29·4 – 50·7) and nursing staff (38·8%, 95% CI 32·1 – 45·7). while consultant, trainee doctors and house officer had seroprevalence of (18·2%, 95% CI 12·4 – 25·1), (19·9%, 95% CI 14·3 – 26·4) and (18·4%, 95% CI 11·7 – 26·7) respectively [Table:1 & Graph:2].

**Graph 2:**
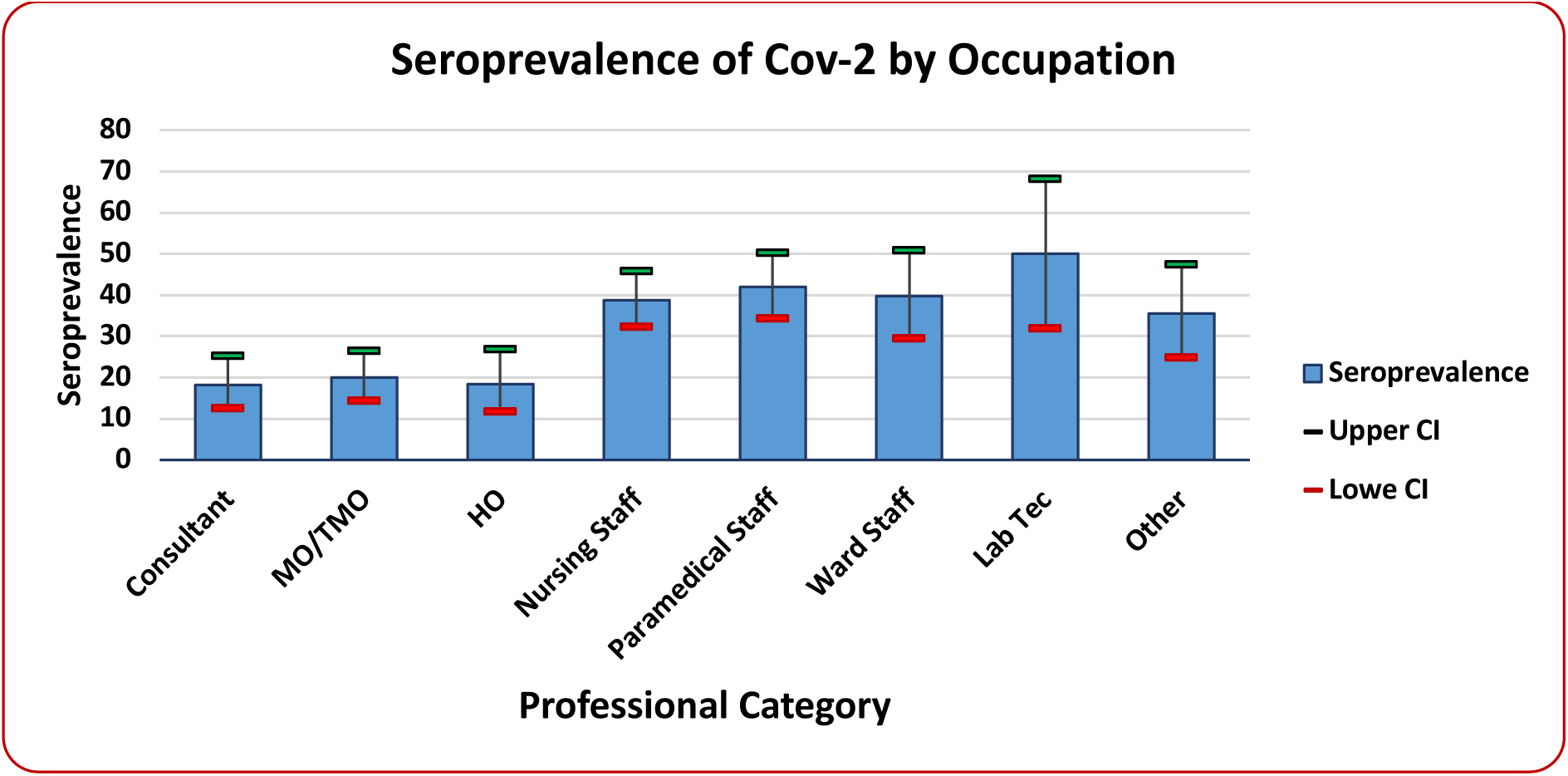

Among the seropositive HCWs, 165 (53·2%) were completely asymptomatic while 145 (46·8%) had one or more symptoms. The mean Antibody level was 26·12 (SD ± 26·79) AU/ml in seropositive participants (Males 24·63 SD ±25·68, Females 29·72 SD ±29·14). The mean antibody level in seropositive asymptomatic participants was 30·20 (SD ± 29·63) while in symptomatic it was 21·48 (SD ± 22·35).

### Risk Factors

Gender was not an independent risk factor and the odds of being seropositive were similar between males and females (OR: 1·02, 95% CI, 0.89-1·41. p> 0·05).

The use of face masks and observing social distancing within a family had lesser odds of being seropositive with a statistical significant association (OR: 0·67, 95% CI, 0·49 – 0·92. p<0·05), (OR: 0·73, 95% CI, 0·55 – 1·98. p<0·05) in multivariable regression models (MLM) [Table: 2]. In MLM, the odds of seropositivity were higher among those attending funeral and visiting local markets for shopping (OR: 1·83, 95% CI, 1·05 – 3·16. p<0·05) and (OR: 1·66, 95% CI, 1·16 – 2·37. p<0·01). However the risk of seropositivity did not increase with attending congregational prayers in mosques (OR: 0·52, 95% CI, 0·34 – 0·79. p<0·05) [Table: 2].

**Table: 2:** Univariable and Multivariable analysis of factors associated with seropositivity.

|  | Univariable Analysis |  |  | Multivariable Analysis |  |  |
| --- | --- | --- | --- | --- | --- | --- |
|  | OR | 95%CI | P value | OR | 95% CI | P value |
| Age | 1.00 | 0.99-1.01 | 0.66 | 1.00 | 0.99-1.01 | 0.71 |
| Gender | 1.19 | 0.89-1.59 | 0.24 | 1.02 | 0.89-1.41 | 0.21 |
| **Professional Category** |  |  |  |  |  |  |
| Consultant | 0.44 | 0.29-0.62 | 0.00* | 0.38 | 0.15-0.97 | 0.04* |
| MOs/TMOs | 0.49 | 0.33-0.73 | 0.00* | 0.42 | 0.17-1.04 | 0.06 |
| HOs | 0.47 | 0.29-0.77 | 0.03* | 0.40 | 1.21-4.04 | 0.67 |
| Nursing Staff | 1.58 | 1.15-2.17 | 0.05* | 1.20 | 1.17-4.65 | 0.68 |
| Paramedical Staff | 1.79 | 1.26-2.53 | 0.01* | 1.12 | 0.43-1.37 | 0.78 |
| Ward Staff | 1.49 | 0.95-2.33 | 0.78 | 1.10 | 0.44-1.39 | 0.82 |
| Lab Technician | 2.26 | 1.27-4.04 | 0.05* | 1.49 | 0.43-1.55 | 0.32 |
| Other | 0.55 | 0.23-1.27 | 0.16 | 1.08 | 0.24-1.33 | 0.85 |
| **Exposure Risk** |  |  |  |  |  |  |
| Observe social distancing | 1.0 | 0.73-1.46 | 0.82 | 1.0 | 0.70- 1.53 | 0.83 |
| Using face mask | 0.69 | 0.51-0.93 | 0.01 | 0.67 | 0.49 – 0.92 | 0.01* |
| Travel abroad | 0.70 | 0.50-0.98 | 0.03 | 0.73 | 0.52 – 1.04 | 0.08 |
| Social distancing with-in Family | 1.29 | 0.99-1.69 | 0.05 | 0.73 | 0.55 - 0.98 | 0.04* |
| Wedding | 0.85 | 0.37-1.95 | 0.71 | 1.0 | 0.41 – 2.45 | 0.98 |
| Social Gathering | 0.81 | 0.38-1.67 | 0.55 | 0.93 | 0.41 – 2.12 | 0.87 |
| Funeral | 1.78 | 1.07-2.96 | 0.02 | 1.83 | 1.05 – 3.16 | 0.03* |
| School/madrassa attending | 1.13 | 0.10-12.5 | 0.92 | 0.94 | 0.80 – 11.2 | 0.96 |
| Local Markets | 1.55 | 1.15-2.08 | 0.00 | 1.66 | 1.16–2.37 | 0.00* |
| Shopping malls | 0.75 | 0.43-1.30 | 0.31 | 1.0 | 0.54 – 1.88 | 0.96 |
| Masjid | 0.55 | 0.37-0.80 | 0.00 | 0.52 | 0.34 – 0.79 | 0.001* |
| Direct contact with COVID patient | 0.72 | 0.54-0.94 | 0.01 | 0.83 | 0.62 – 1.11 | 0.22 |
| Contact with COVID patient outside hospital | 1.0 | 0.78-1.66 | 0.72 | 1.0 | 0.63 – 1.53 | 0.87 |
| **Symptoms** |  |  |  |  |  |  |
| Fever | 2.14 | 1.54-2.97 | 0.00 | 2.36 | 1.52-3.68 | 0.00* |
| Cough | 1.72 | 1.23-2.41 | 0.00 | 1.44 | 0.93-2.24 | 0.10 |
| Sneezing | 0.17 | 0.51-1.12 | 0.17 | 0.56 | 0.63-1.12 | 0.10 |
| Sore Throat | 0.79 | 0.57-1.10 | 0.17 | 0.78 | 0.57-1.11 | 0.16 |
| Breathlessness | 2.13 | 0.96-4.72 | 0.06 | 1.31 | 0.52-3.25 | 0.56 |
| Body Aches | 1.0 | 0.79-1.50 | 0.58 | 0.80 | 0.53-1.19 | 0.27 |
| Diarrhea | 1.39 | 0.89-2.17 | 0.14 | 1.26 | 0.72-2.20 | 0.40 |
| Loss of taste | 2.40 | 1.44-4.00 | 0.00 | 1.24 | 0.62-2.50 | 0.53 |
| Loss of smell | 3.80 | 2.18-6.61 | 0.00 | 2.95 | 1.46-5.98 | 0.00* |
| Pain Abdomen | 0.72 | 0.39-1.34 | 0.30 | 0.56 | 0.27-1.16 | 0.12 |
| Severe headache | 1.0 | 0.64-1.59 | 0.95 | 0.77 | 0.44-1.38 | 0.39 |

Seroprevalence in different professional category ranged from 18·2% (95% CI, 12·4-25·1) in consultants to 50.0% (95% CI, 31.8-68.1) in laboratory technicians. In Univariable analysis, being a nursing or paramedical staff led to higher risk of seropositivity (OR, 1·58, 95% CI, 1·15-2·17. p< 0·05),(OR, 1·79, 95% CI, 1·26-2·53. p< 0·05) but multivariable logistic regression did not show any significant association [Table: 2].

The risk of being seropositivity was strongly (p< 0.01) associated with fever (OR: 2·36, 95% CI: 1·52–3·68) and loss of smell (OR: 2·95, 95% CI: 1·46–5·98) while loss of taste was strongly associated with seropositivity (OR: 2·4, 95%CI, 1·44-4.00, p<0·001) in univariable analysis but multivariable logistic regression did not show any significant association. [Table: 2] Co-morbidities were present in 17% in seropositive subjects and included diabetes (30%), hypertension (36·4%), cardiac disease (15·4%), asthma (18·2%) and recent surgery (40%).

## Discussion

To our knowledge this is the first study on prevalence of SARS CoV-2 antibodies in HCW of tertiary care hospitals in Pakistan. Studies form other counties observed lower seroprevalence in HCWs. The seroprevalence of SARS-CoV-2 antibodies in healthcare workers were 30.7% (CI 95%: 27·8 – 33·6). It varied from 18.2% among doctors to 50% in laboratory technicians. The highest seroprevalence were reported in Lab technicians (50%) and paramedical staff (42%) compared to the rest of HCWs. In a study from China, the seroprevalence was 17.14% while 24% and 9·3% have been reported from UK and Spain.^12,13,14^ Much lower weighted prevalence (1·07%) was reported in a Greek study in 1952 HCWs.^15^

The higher seroprevalence of antibodies in our study may indicate higher exposure of HCWs to COVID-19 positive subjects or patients. It may also be due to inadequate use of PPE and education/awareness levels of HCWs. A recent meta-analysis published in the Lancet Journal concluded that physical distancing, use of mask and goggles significantly decrease the risk of infection.^16^ However even in developed countries there have been problems with adequate supplies of PPE and 65% resident physicians in New York considered it inadequate as reported by the program director.^17^ This warrants for adequate provision of PPE and better training and awareness of HCWs against COVID-19 in HCWs working in tertiary care hospitals in Pakistan.

The risk of seropositivity was significantly high in subject with history of attending funerals (OR:1·83 95% CI, 1·05-3·16) and visiting local markets (OR: 1·66, 95% CI, 1·16-2·37) but not in subjects attaining shopping malls. This difference in risk may be due to better observance of preventive protocols, provision of sanitizers at the entrances of shopping malls, where the clients are usually from middle and high income class of the society. In contrast the local markets consist of clusters of small shops providing commodities of common use at cheaper rates. The clients are usually low income group people and there are multiple open accesses of such markets, sanitizers are not provided and social distancing is not observed.

The risk of increased seropositivity was also not associated with attending congregational prayers in mosques (OR:0·52, 95% CI, 0·34-0·79). This could be possibly due to two main reasons. First, the overall personal and environmental hygienic practices observed as religious obligation in mosques that includes washing hands and face at least five times a day before prayers and keeping the prayer area clean. Second, voluntary implementation of preventive measures after the consensus decrees on the same by religious scholars.^18^ This also highlights the need of involvement of clergy for effective implementation of public health strategies in conservative societies like Pakistan.

The risk of becoming positive for SARS-CoV-2 antibodies did not increase with history of direct contact with COVID patients within or outside the hospital. This could be due to more careful approach of HCWs when coming in contact with known COVID patients. The same has been reported in other studies that frontline HCWs dealing with COVID patients do not show higher risk of acquiring the infection when compared to Non frontline HCWs.^19^

In our study most of the subjects were asymptomatic. The mean antibodies level in Seropositive asymptomatic participants were significantly higher compared to symptomatic subjects (p<0·001). In contrast other studies reported lower antibodies level in asymptomatic patients.^20^ It is also suggested that asymptomatic patients may have lower seroconversion levels but the duration of virus shedding is longer in them when compared to symptomatic patients^21^. The asymptomatic HCWs could therefore be potential threat of transmitting infection.

The large number of asymptomatic HCWs in our study could have been potential source of transmission of infection to their colleagues, hospital staff and contacts outside hospital. This could be one reasons of higher exposure of HCWs to SARS CoV-2 virus and consequently higher seroprevalence in this study. The HCWs need to be more aware of this problem and should preferably have lower threshold of screening themselves with subtle symptoms or even no symptoms. The management of hospitals may offer optional “spot” screening to HCWs. The proposed strategy might pick up otherwise unidentified positive cases for further workup and help in adopting appropriate preventive measures to reduce the transmission of infection.

In multivariate regression analysis two major symptoms i.e. history of fever [OR: 2·36, 96% CI, 1·52-3·68, p<0·001] and loss of smell [OR: 2·95, 95% CI, 1·46-5·98, p<0.001] were strongly associated with seropositivity. Others symptoms including cough, breathlessness and loss of taste showed a significant correlation in univariate analysis but multivariate analysis did not show a significant association.

The risk of becoming seropositive was not different significantly in males and females but the mean antibodies titres were significantly high in females (P<0·03).

Increasing age was a significant risk for SARS-CoV 2 antibodies levels. The highest mean antibody level (38·95 ± 34·88) was seen in the age group (50-59) while the lowest (23·73 ± 25·44) was in the age group (20-29) (p = 0·05). In a mathematical model to epidemic data from six countries a positive correlation was found with increasing age and susceptibility of young was almost half to that of adults.^22^

Profession of HCWs was a significant risk and seropositivity with higher prevalence in nursing and paramedical staff compared to consultants and trainee doctors (HOs and MOs/TMOs) in univariate analysis. This is consistent with SARS-CoV study epidemic in 2003.^23^ and could be due to longer duration of contact (more than 30 minutes) ^14^ of specific HCWs. However multivariate analysis did not show any significant difference.

The three commonest reported co-morbidities in other studies are hypertension, diabetes and cardiovascular diseases.^24^ In our study the overall co-morbidities were 17% in seropositive subjects and these were recent history of surgery 40%, hypertension 36·4%, diabetes 30%, asthma 18·2% and cardiac disease 15·4%.

## Conclusions

The HCWs in our setup are at high risk of acquiring COVID-19 infection. They need better education on risk factors, training and possibly adequate provision of PPEs to reduce the risk of infection. A large number of asymptomatic HCWS could be potential threat of transmitting infection to others. The HCWs should have a lower threshold for screening of COVID-19 to pick up positive cases and reduce the potential risk to others. SARS Co V 2 antibodies positive HCWs may be considered for voluntary deployment in COVID caring hospitals as they may be at lower risk of acquiring COVID infection. The clergy should be involved in effective implementation of public health strategies in conservative societies like Pakistan.

## Data Availability

All data related to our research study is avilable and can be provided upon request

## Notes

### Competing Interest Statement

The authors have declared no competing interest.

### Funding Statement

Funding Agency: Prime Foundation Pakistan
Grand no: 2020-6-CG-0001

### Author Declarations

Ethical approval obtained from Ethical Review board of Prime Foundation, Pakistan

